# Low awareness of past SARS-CoV-2 infection in healthy adults

**DOI:** 10.1101/2020.08.10.20171561

**Authors:** Katja van den Hurk, Eva-Maria Merz, Femmeke J. Prinsze, Marloes L.C. Spekman, Franke A. Quee, Steven Ramondt, Ed Slot, Hans Vrielink, Elisabeth M.J. Huis in ’t Veld, Hans L. Zaaijer, Boris M. Hogema

## Abstract

**Background:** The coronavirus disease 2019 (COVID-19) pandemic challenges governments worldwide to balance appropriate virus control measures and their societal and economic consequences. These control measures include the identification, isolation and testing of potentially infected individuals. As this relies on an individual’s awareness of infection, we investigated the extent to which healthy adults suspected having had COVID-19, and how COVID-19 suspicion and symptoms relate to antibodies indicative of a past infection with the severe acute respiratory syndrome coronavirus 2 (SARS-CoV-2).

**Methods and findings:** For this cross-sectional study, individuals donating plasma anywhere in the Netherlands between May 11^th^ and 18^th^ were screened for total SARS-CoV-2 antibodies using ELISA and invited to participate in an online questionnaire about COVID-19-related symptoms and awareness. Antibody and questionnaire data were complete for 3,676 individuals, including 239 (6.5%) that tested positive for SARS-CoV-2 antibodies. Here, we show that a 38% of the individuals that tested positive for SARS-CoV-2 antibodies reported having had no or only very mild symptoms at any time during the peak of the epidemic. The loss of taste and/or smell in particular was significantly associated with seropositivity, independent of age and sex. Forty-eight percent of antibody-positive persons did not suspect having had COVID-19, in spite of most of them reporting symptoms.

**Conclusions:** Awareness of infection was low among individuals that tested positive for SARS-CoV-2 antibodies, even at the peak of the epidemic. Improved awareness and recognition of COVID-19 symptoms and tracing of asymptomatic contacts is crucial to halting SARS-CoV-2 transmission.

## Introduction

Due to the current coronavirus disease (COVID-19) pandemic caused by the severe acute respiratory syndrome coronavirus 2 (SARS-CoV-2), governments worldwide are struggling to find an appropriate balance between virus control measures and their societal and economic consequences.^1^ Physical distancing and (partial) closures of offices, nursing homes, restaurants, schools and shops have played – and still play – an important role in combating the spread of SARS-CoV-2. An impending economic crisis and the huge societal burden call for informed easing of these measures.

Currently limited knowledge exists regarding the extent to which SARS-CoV-2 infections may remain undetected, while pre- and asymptomatic individuals are thought to contribute significantly to the spread of SARS-CoV-2.^2,3^ A wide clinical spectrum of SARS-CoV-2 infections has been described, ranging from mild flu-like symptoms to severe viral pneumonia with respiratory failure, and death.^4,5^ Due to the limited availability of tests and infrastructure, more severe COVID-19 cases are likely overrepresented in the majority of studies conducted thus far. Many cases may remain undetected in the event of asymptomatic infection, mild infection with isolated symptoms such as olfactory loss, or symptomatic infections that are attributed to other causes.^3,6,7^

Post-lockdown measures often rely on individuals, in particular those who have been in contact with a confirmed COVID-19 case, to self-isolate and get tested in the event of COVID-19-related symptoms. These measures are dependent on individuals’ recognition of symptoms, yet it is unknown whether or not infected individuals are able to identify themselves as such. Hence, we studied the association between COVID-19 suspicion and SARS-CoV-2 antibody status, as well as that between self-reported symptoms and antibody status in healthy adults.

## Methods

### Study population

The first case of COVID-19 in the Netherlands – one of the most densely populated countries in Europe with 17.2 million inhabitants – was reported on February 27^th^, 2020. By the 11^th^ of May, 42,788 PCR-confirmed COVID-19 cases were reported, including 11,343 hospital admissions and 5,456 deaths.^8^ Disease prevalence showed a positive North-South gradient.

Sanquin is by law the only organization authorized to collect and distribute blood and blood components in the Netherlands. On a yearly basis, approximately 330,000 individuals aged 18 to 73 years make over 700,000 donations on a voluntary non-remunerated basis. Of those donations, 300,000 are apheresis procedures, which separate specific blood components, such as plasma, and return the remainder to the donor. Individuals must be in good health to be eligible to donate, which is checked using a pre-donation donor health questionnaire and capillary haemoglobin measurement.^9^ Additional screening at the donation centres currently prevents individuals with any potential COVID-19-related symptoms from donating.

Individuals that donated plasma anywhere in the Netherlands between May 11^th^ and 18^th^ 2020 who consented (99.7%) to using leftovers of their donation for research were tested for SARS-CoV-2 antibodies. Those that donated COVID-19 convalescent plasma were excluded, as they were recruited separately and are, thus, not representative of regular plasma donors. Eligible individuals with registered e-mail addresses were subsequently invited to participate in an online questionnaire. This cross-sectional study was conducted according to the principles of the Declaration of Helsinki (2013) and the General Data Protection Regulation (GDPR). All participating individuals gave informed consent before participating in the online questionnaire and the study protocol and procedures were approved by Sanquin’s Ethics Advisory Council and its Privacy Officer. The datasets generated and/or analyzed in the context of this study are available from the corresponding author upon reasonable request.

### SARS-CoV-2 antibody testing

Plasma samples were tested for the presence and levels of antibodies against SARS-CoV-2, using a SARS-CoV-2 total antibody ELISA (Wantai Biological Pharmacy Enterprise Co., Beijing, China) following the manufacturer’s instructions. Samples that tested reactive, including samples with weak reactivity (OD/CO ratio ≥0.5), were re-tested and considered positive if the re-test was reactive (OD/CO ratio >1.0). We previously determined the sensitivity and specificity of the Wantai test to be 98.7% and 99.6%.^10^ As OD/CO ratios were not normally distributed, antibody levels were dichotomized (>10.0 considered high).

### Online questionnaire

Within 8 days after their donation, all individuals for whom e-mail addresses (n=7,721) were available and valid were invited by e-mail and provided with a web link to an online questionnaire, programmed in Qualtrics (SAP, Walldorf, Germany). The questionnaire included self-reported COVID-19 status (diagnosed by PCR/suspected/not suspected) and a list of 18 symptoms considered to be COVID-19-related according guidelines specified by the Dutch National Institute for Public Health and the Environment.^11^ Participants indicated the extent to which they suffered from these symptoms in the period between one week before the first confirmed case nationally (February 21^st^, 2020) and their donation date on a scale from 0 (not at all) to 5 (severely). A symptom was considered present if the score was 1 (very mild) or higher. Symptom severity was defined as asymptomatic (score 0), only very mild symptoms (score 1) or mild to severe symptoms (score for at least one symptom 2 or higher). Participants that reported symptoms were also asked whether or not they consulted a physician or were admitted to a hospital and what they thought had caused their symptoms: temporary illness (e.g. flu, COVID-19), chronic complaints, other circumstances (e.g. allergies, trauma) or unknown.

Demographic background data on age, sex, and region were obtained from the blood bank information system eProgesa (MAK systems, Paris, France).

### Statistical analyses

Descriptive information for continuous variables was calculated as mean and standard deviations, or median and interquartile range if skewed. We performed two-sided t-tests for continuous variables and chi^2^ tests for proportions to assess differences between subgroups. Associations between symptoms and disease severity with antibody status and levels were estimated using logistic regression analyses. Statistical analyses were performed using SPSS, Version 23 (IBM, Armonk, U.S.A.).

## Results

Of the 8,275 donors that underwent plasmapheresis between 11^th^ and 18^th^, we tested 7,150 for SARS-CoV-2 antibodies, of whom 419 (5.9%) tested positive. OD/CO ratio in seropositive individuals ranged from 1.01 to 20.86. We invited 7,721 individuals to participate in the online questionnaire, of whom 4,275 (55.4%) participated. Antibody and questionnaire data were complete for 3,676 individuals, including 239 (6.5%) who tested positive for SARS-CoV-2 antibodies. Seropositive individuals were generally younger and more likely to live in the Southern region of the Netherlands than seronegative individuals (Table 1). Forty-eight percent of the seropositive individuals, and 87% of those seronegative, did not suspect they had had COVID-19. About 11% of the seropositive individuals reported no symptoms at all. An additional 27% of seropositive individuals reported only very mild symptoms, generally sneezing (69%), a cold (55%), and/or fatigue (40%). Only one individual positive for SARS-CoV-2 antibodies was admitted to a hospital, but this was because of gastrointestinal complaints.

**Table 1.** Characteristics and COVID-19 status, stratified by SARS-CoV-2 antibody status.

|  |  | **SARS-CoV-2**<br>**antibody**<br>**positive** | **SARS-CoV-2**<br>**antibody**<br>**negative** | **P values** |
| --- | --- | --- | --- | --- |
| **Total** |  | 239 | 3,437 |  |
| Sex | Male | 126 (52.7%) | 1,766 (51.4%) | 0.696 |
|  | Female | 113 (47.3%) | 1,671 (48.6%) |  |
| Age (in years) |  | 46.6 ± 13.8 | 50.0 ± 14.0 | <0.001 |
| Region* | South | 98 (41%) | 943 (27%) | <0.001 |
|  | Mid | 114 (48%) | 1,846 (54%) |  |
|  | North | 27 (11%) | 641 (19%) |  |
| COVID-19 status | Infection diagnosed** | 3 (1.3%) | 0 (0.0%) | <0.001 |
|  | Infection suspected | 125 (52.3%) | 435 (12.7%) |  |
|  | Infection not suspected | 114 (47.7%) | 3,002 (87.3%) |  |
| Symptom severity | Asymptomatic | 26 (10.9%) | 1,047 (30.5%) | <0.001 |
|  | Only very mild symptoms | 64 (26.8%) | 1,250 (36.4%) |  |
|  | Mild to severe symptoms | 149 (62.3%) | 1,134 (33.0%) |  |
| Contacted physician |  | 30 (12.6%) | 270 (7.9%) | 0.009 |
| Admitted to hospital |  | 1 (0.4%) | 14 (0.4%) | 0.980 |

Numbers (% within antibody status) and mean ± standard deviations are shown. P values indicate the significance of differences; a 2-sided t-test for age, $\chi^2$ tests for proportions. \*North=provinces of Friesland, Groningen, Drenthe and Overijssel, South=North Brabant, Limburg and Zeeland, Mid=all other provinces. \*\*Self-reported PCR-confirmed diagnoses.

Apart from diarrhoea, regurgitation, rash, and confusion, all symptoms were significantly more frequently reported by seropositive versus –negative individuals (Figure 1). Anosmia/dysgeusia was the symptom most strongly associated with the presence of SARS-CoV-2 antibodies; the odds ratio of 12.7 was significantly higher than for any other COVID-19-related symptom (Table 2). Despite this strong association, it was not the most prevalent symptom among seropositive individuals. Other symptoms, such as fever, a cold, fatigue, coughing, headaches, muscle and joint pains, and sneezing were similarly or even more prevalent among these individuals. However, these symptoms appeared less indicative of a SARS-CoV-2 infection.

**Figure 1.**
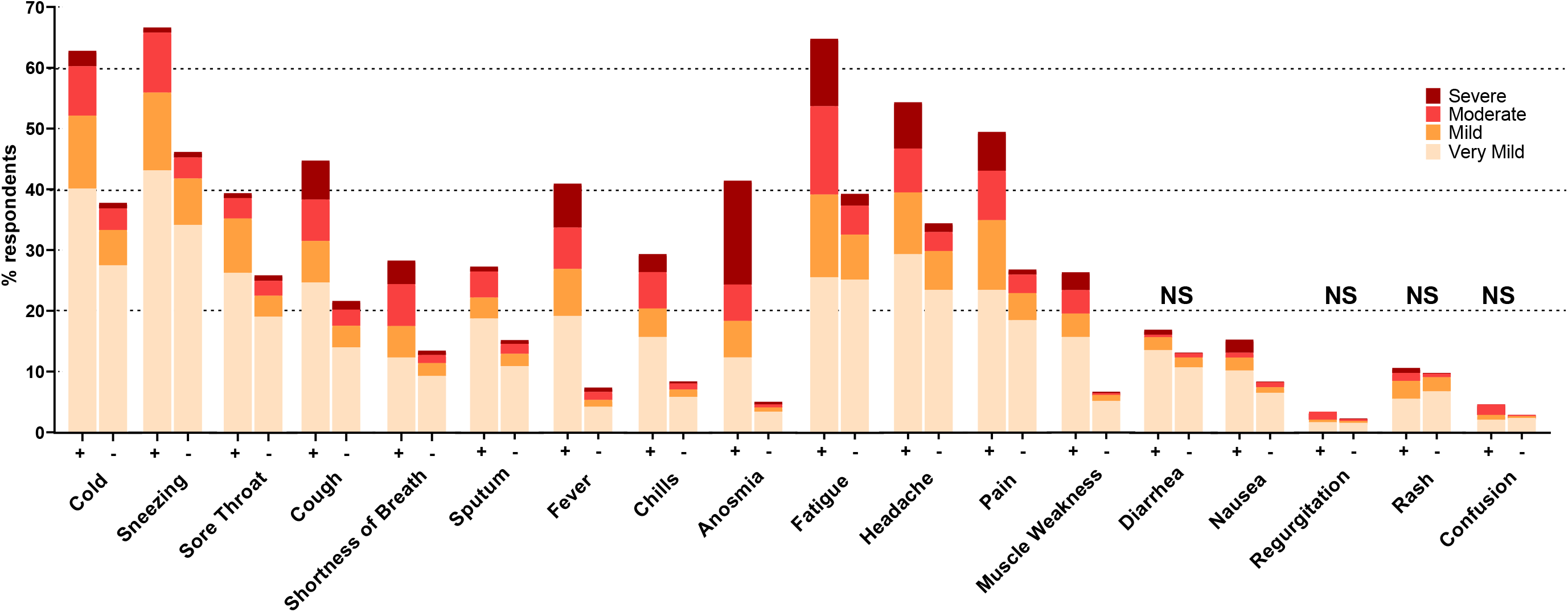
Percentages of individuals that reported very mild to severe symptoms by antibody status: positive (+) or negative (-). Age and sex-adjusted logistic regression models indicate higher prevalence in antibody positive versus -negative individuals for all symptoms (p<0.001, Table 2), except where indicated as not significant (NS). Anosmia refers to anosmia and/or dysgeusia, pain refers to muscle and/or joint pain.

**Table 2:**
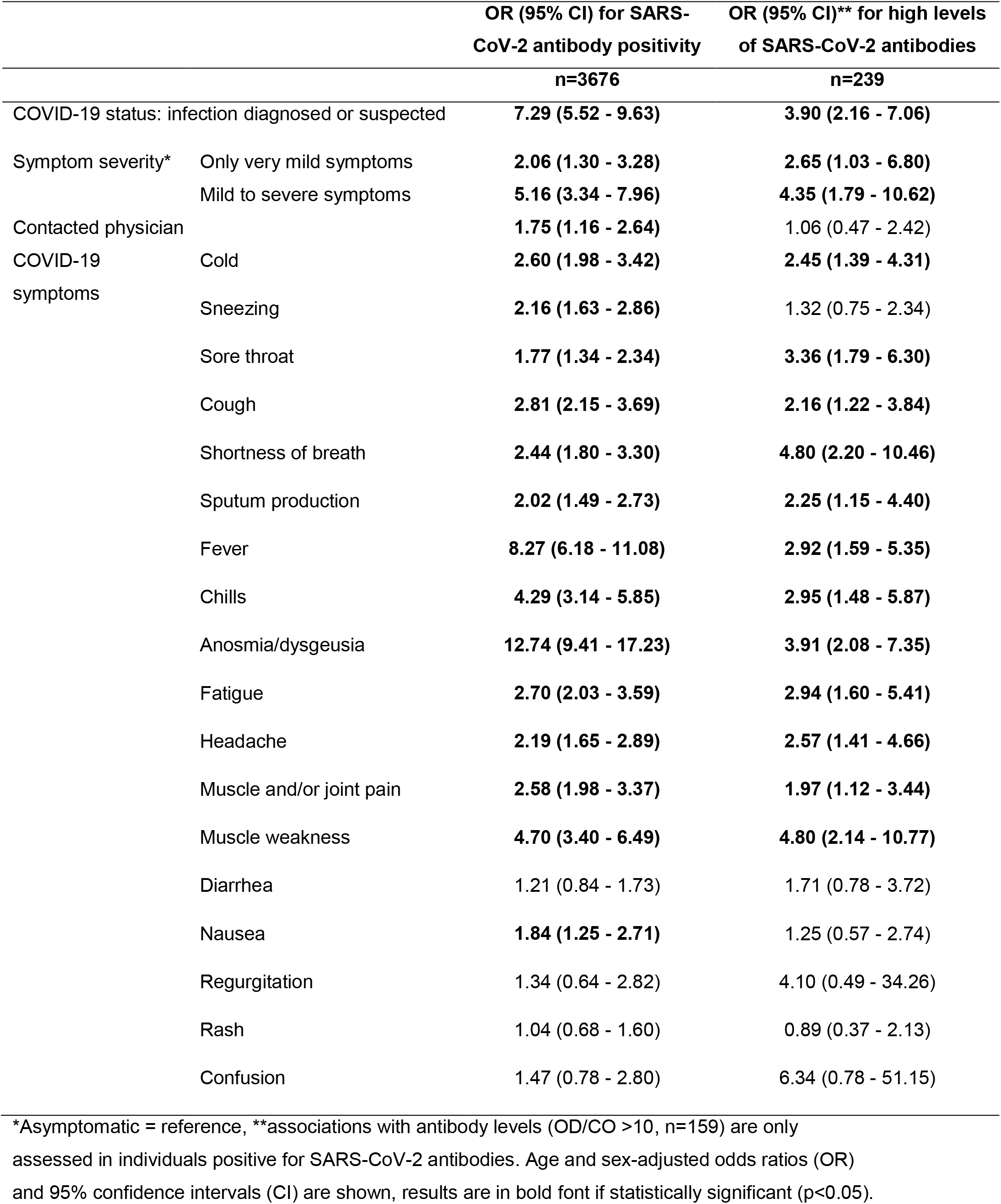
Associations between COVID-19 suspicion, symptom severity and symptoms with SARS-CoV-2 antibody positivity and high levels of SARS-CoV-2 antibodies.

In seropositive individuals, the presence of symptoms was also significantly associated with high levels of SARS-CoV-2 antibodies, except for the symptoms sneezing and nausea (Table 2). Shortness of breath and muscle weakness were most strongly associated with high levels of SARS-CoV-2 antibodies. Individuals that suspected having had a SARS-CoV-2 infection were significantly more likely to be SARS-CoV-2 antibody positive and, among those antibody-positive individuals, to have high levels of antibodies compared to individuals that did not suspect having had COVID-19. More severe symptoms versus being asymptomatic was also associated with antibody status and levels, as was consultation of a physician because of those symptoms.

Of all individuals that tested positive for SARS-CoV-2 antibodies, those who did not suspect having had the infection reported no or very mild symptoms in 19 and 41% of all cases, compared to only 3 and 14%, respectively, of those who did (Table 3). Physician contact and hospital admittance did not differ between antibody-positive individuals who did or did not suspect having had COVID-19. Individuals that did not suspect having had COVID-19 attributed their symptoms to unrelated circumstances, such as allergies in 39% of the cases, or temporary illness such as the flu, unknown reasons or chronic complaints in 20, 18 and 4% of the cases, respectively.

**Table 3.** Symptom severity, health care use and suspected causes of symptoms, stratified by COVID-19 suspicion in individuals positive for SARS-CoV-2 antibodies.

|  |  | COVID-19<br>diagnosed or<br>suspected | COVID-19 not<br>suspected | P values |
| --- | --- | --- | --- | --- |
|  |  | n=125 | n=114 |  |
| Symptom severity | Asymptomatic | 4 (3.2%) | 22 (19.3%) | <0.001 |
|  | Only very mild symptoms | 17 (13.6%) | 47 (41.2%) |  |
|  | Mild to severe symptoms | 104 (83.2%) | 45 (39.5%) |  |
| Contacted a physician |  | 15 (12.0%) | 15 (13.2%) | 0.947 |
| Admitted to hospital |  | 0 (0.0%) | 1 (0.9%) | 0.477 |
| Likely cause of<br>symptoms (if any) | Temporary illness | 106 (84.8%) | 23 (20.1%) | <0.001 |
|  | Chronic complaints | 0 (0.0%) | 4 (3.5%) |  |
|  | Unrelated circumstances* | 5 (4.0%) | 44 (38.6%) |  |
|  | Unknown | 10 (8.0%) | 20 (17.5%) |  |

\*Circumstances not related to COVID-19, such as allergies or trauma. Numbers (% within COVID-19 suspicion status) are shown, differences tested with Chi<sup>2</sup>, or Fisher's exact in the event of cells with expected counts less than 5.

## Discussion

We explored associations between COVID-19 suspicion and SARS-CoV-2 antibody status, as well as between symptoms and antibody status in healthy adults. Of those with reactive test results, 48% did not suspect having been infected with SARS-CoV-2. Eleven percent reported a complete absence of symptoms and 27% only very mild symptoms during the national peak of the epidemic. COVID-19-related symptoms – particularly anosmia/dysgeusia and fever – were significantly associated with antibody status, independent of age and sex.

To the best of our knowledge, we are the first to show a lack of COVID-19 suspicion in almost half of the subgroup that tested positive for SARS-CoV-2 antibodies. This may have an impact on individual adherence to governmental measures and on the decision to request a PCR test, as health behavior largely depends on beliefs regarding one’s own and others’ perceived susceptibility to and severity of disease.^12^ A behavioral study by the National Institute for Public Health and the Environment showed that 80% of the people that reported symptoms did not stay inside their homes and 40% even went to work.^13^ Also, while 68% of the study participants indicated that they will get tested if they develop symptoms, this percentage drops to 28% among individuals once they displayed symptoms. The main reason for this was that individuals attributed their symptoms to hay fever or a common cold, of which the latter may occur even more frequently during the flu season.

Assuming that the vast majority of SARS-CoV-2 infections induce an antibody response, 38% of the infected individuals reported having no (11%) or only very mild symptoms (27%).^14^ An important advantage of our retrospective study design in comparison to previous studies reporting 40-45% asymptomatic infections is the unlikelihood of falsely identifying pre-symptomatic cases as asymptomatic.^3^ In addition, our thorough assessment of symptoms over an extended period is likely to have captured milder symptoms that may have been underreported in previous studies. Indeed, previous studies have been mainly cross-sectional or with incomplete follow-up and with register-based symptoms rather than systematic assessments.^15,16^

Particularly those individuals that reported no or very mild symptoms only were relatively unlikely to suspect having been infected with SARS-CoV-2. On the other hand, our study shows that in comparison to being asymptomatic, even very mild symptoms, and mild to severe symptoms in particular, were associated with SARS-CoV-2 infection, as well as with high levels of SARS-CoV-2 antibodies. Our finding that anosmia/dysgeusia is the symptom most strongly associated with COVID-19 is in line with results from the COVID Symptom Study, which showed this symptom to be the strongest predictor of PCR-confirmed SARS-CoV-2 infection.^17^ Improved awareness and recognition of COVID-19 symptoms, in particular of the loss of smell and taste, may therefore help to reduce the proportion of undetected COVID-19 cases.

Strengths of this study include the large study sample, the superior performance of the Wantai SARS-CoV-2 total antibody ELISA over other antibody tests, and the thorough questionnaire-based assessment of the presence and severity of symptoms.^18^ Our study also has limitations. First, study participants were required to be in good health in order to qualify for plasma donation. This selection bias may have resulted in an underrepresentation of SARS-CoV-2 antibody positive individuals and of more severe COVID-19 cases in particular. Second, we asked participating individuals to report their symptoms over a period of almost three months, which obviously introduces a risk of recall bias. Nonetheless, given the unprecedented impact of SARS-CoV-2 on society and the repeated governmental calls to self-isolate with symptoms, we expect that most individuals have an exceptional recollection of their COVID-19 suspicion and symptoms during this period.

As governments slowly ease virus control measures, it becomes vital to identify and isolate infected individuals to prevent new SARS-CoV-2 outbreaks.^19,20^ The presence of anosmia/dysgeusia, especially, should trigger PCR testing.^17^ In addition, our study confirms the existence of asymptomatic SARS-CoV-2 infections and adds that even symptomatic individuals did not suspect a SARS-CoV-2 infection. Despite the limitations of studies thus far, sufficient evidence of asymptomatic and pre-symptomatic transmission of SARS-CoV-2 exists.^16,21^ Efforts to identify cases that rely on symptoms may therefore be insufficient, which emphasizes the importance of thorough contact tracing.^22^

In conclusion, almost half of the individuals that tested positive for antibodies to SARS-CoV-2 in a reliable assay did not suspect having had an infection. This proportion may be lowered with better awareness and recognition of COVID-19 symptoms, in particular the loss of smell and taste. However, 38% of those infected reported no or only very mild symptoms. Tracing of asymptomatic contacts is therefore crucial to halting transmission.

## Data Availability

The datasets generated and/or analyzed in the context of this study are available from the corresponding author upon reasonable request.

## Author contributions

KvdH and FJP analysed data, FJP, MLCS, FAQ, SR and HV provided project support and organized participant recruitment, KvdH, EMM, ES, EMJHitV, HLZ, and BMH designed and ran the study and wrote the manuscript. All authors critically revised and approved the final version of the manuscript.

## Code availability

The computer code used to generate results that are reported in this paper are available to editors and reviewers upon request.

